# Is a 24-Month Birth Interval Enough? Evidence from a cross-sectional study using the Benin Demographic and Health Survey

**DOI:** 10.64898/2026.07.16.26358222

**Authors:** Mahugnon Christian Urlyss Agossou, Giulia Scarpa, Lenka Beňová, Christelle Boyi Hounsou, Diana Sagastume, Gottfried Agballa, Jean-Paul Dossou, Kerry LM Wong

## Abstract

**Background:** Stunting affects approximately 32% of children under five years in Benin. While birth intervals shorter than 33 months are a recognized risk factor for childhood malnutrition, the optimal birth interval for preventing stunting in the Beninese context is still unclear.

**Objective:** This study examined the association between preceding birth interval (PBI) and stunting among children aged 6-59 months in Benin.

**Methods:** This study used a cross-sectional design to analyze data from the 2017-18 Benin Demographic and Health Survey. We included 10,153 children aged 6-59 months. Stunting was defined as height-for-age z-score below 2 standard deviations from World Health Organization standards. Preceding birth interval was categorized as <24, 24-32, 33-44, 45-56, and >56 months. Survey-adjusted multivariable logistic regression was used to estimate adjusted odds ratios (aORs) and 95% confidence intervals for the association between PBI and stunting, controlling for child, maternal, and household-level covariates. Potential effect modification by child age group (6–23 vs. 24–59 months) was assessed through a multiplicative interaction term and evaluated using information criteria, a likelihood-ratio test, and the statistical significance of individual interaction terms.

**Results:** Compared with children born after an interval of <24 months, those born after 45–56 months (AOR: 0.63; 95%CI: 0.51–0.78) and >56 months (AOR: 0.67; 95%CI: 0.54–0.82) had significantly lower odds of stunting (both p<0.001). Intervals of 24–32 months and 33–44 months were not significantly associated with stunting, nor was first-born status. No evidence of effect modification by child age group was found (likelihood-ratio test p=0.336), and stratified analyses conducted separately for children aged 6–23 months and 24–59 months yielded results consistent with those from the pooled model.

**Conclusion:** The lack of protective effect for intervals shorter than 45 months indicates a context-specific threshold above which nutritional benefits become manifest. Integrating family planning messages emphasizing birth intervals longer than 45 months into child nutrition programs, coupled with strengthened access to modern contraception, could contribute to stunting reduction in Benin among other factors. Alongside this, nutritional support for pregnant and breastfeeding women, including adequate dietary supplementation and counselling, may further contribute to improved child growth outcomes

**STRENGTHS AND LIMITATIONS OF THIS STUDY:**

- This study draws on a nationally representative, large-sample dataset (n = 10,153 children aged 6–59 months) collected using internationally standardised methods, ensuring data reliability and generalisability.
- Analyses were adjusted for a comprehensive set of confounders at the child, maternal, and household levels, and stratified by age group (6–23 vs. 24–59 months) to capture age-specific patterns in the association between birth interval and stunting.
- Residual confounding from unmeasured determinants of stunting — including breastfeeding exclusivity, dietary diversity, recurrent infections, and genetic factors — cannot be ruled out despite adjustment for available proxy indicators.
- The data predate recent contextual changes in Benin (e.g., expanded social protection, COVID-19 pandemic, climate-related food insecurity), which may limit the contemporaneous applicability of the findings.

## Introduction

Globally, 23.2% of children under 5 years were stunted in 2024 [1]. While Africa hosts about 32.3% of children under age 5 [2], 43% of those stunted live in Africa [1]. Stunting (defined as being too short for age) reflects chronic undernutrition. It is associated with a higher risk of morbidity and mortality from infections [3–5], sepsis, meningitis, tuberculosis, and hepatitis [5]. Studies conducted in low and middle income countries show stunting before age 2–3 years is associated with poorer cognitive and educational outcomes in later childhood [6–8]. Stunting is associated with multiple maternal, neonatal, child, and environmental factors, including i) at maternal level - short stature, underweight, anemia, infections during pregnancy, and adolescent pregnancy [9]; ii) at neonatal level - fetal growth restriction, preterm birth, and low birth weight [4,9]; iii) at child level - inadequate dietary intake, suboptimal breastfeeding practices, and recurrent infections, particularly diarrhea [4,8,9]; vi) at environmental level - poor sanitation and unsafe water [4,8,9].

In Benin, the prevalence of stunting was similar in 2017 (32%) to that in 1996 (33%), with a peak of 43% in 2006 [10]. Since 2016, the government launched multiple initiatives—such as the National nutrition policy, promotion of exclusive breastfeeding, and integration of nutrition services into primary health care—to accelerate child malnutrition reduction [11]. The country has also joined nutrition-related initiatives, launched several nutrition projects and adopted nutrition strategies, such as the Comprehensive Africa agriculture development program, the Scaling up nutrition movement, the Renewed efforts against child hunger and undernutrition initiative, and the Global alliance for resilience initiative to strengthen governance in nutrition [12]. However, the prevalence of stunting in 2022 is still high at 36.5% in Benin [13].

Beyond the nutritional and environmental factors, birth interval—the time between successive live births—has been identified as a driver of child growth [8,9,14,15]. Biologically, a short birth interval reduces the time for maternal nutritional recovery and may lead to fetal growth restriction and low birth weight [16]. Socially and economically, closely spaced children often compete for parental attention, household resources such as food, and health care [16]. The World Health Organization (WHO) recommended at least 24 months between a live birth to the next pregnancy [17]; this interpregnancy interval corresponds approximately to a minimum birth interval of 33 months. Evidence from a systematic review and meta-analysis links a live birth to the next pregnancy intervals shorter than 24 months (i.e., birth intervals shorter than about 33 months) with higher risk of low birth weight, preterm birth, preterm pre-labor rupture of membranes, intra uterine growth retardation, and perinatal mortality in Sub-Sahara Africa [18].

However, accumulating evidence suggests that these recommendations may not be sufficient across all settings. For example, a national cohort study in Sweden among women with a vaginal first birth found that an interpregnancy intervals shorter than 24–29 months were associated with decreased maternal morbidity and no change in neonatal morbidity, whereas longer interpregnancy intervals were linked to increased risks for both [19]. Research focusing on child nutritional outcomes conversely suggests potential benefits of longer birth spacing in low-income countries [18]. For instance, a 2023 meta-analysis indicated that longer birth intervals—particularly in the range of 36 to 48 months—may be associated with a lower risk of undernutrition among children under five [14]. Together, these findings highlight the need to move beyond a single universal threshold and toward a more context-sensitive understanding of optimal birth spacing [20].

Research specifically examining the relationship between birth interval and stunting in Benin remains limited [21], and it is unclear whether a preceding birth interval (PBI) threshold of 33 months is appropriate in the Beninese context given the country’s nutritional, epidemiological, and socioeconomic profile.

Objective and hypothesis: In this study, we aimed to assess the association between PBI and stunting while adjusting for maternal, child, and household-level factors and using the most recent Benin Demographic and Health Survey data. We hypothesized that a short PBI is associated with a higher odds of stunting.

## Materials and Methods

### Study setting

Benin is a West African country and had a population of 11.9 million inhabitants in 2018, with children under five years accounting for 17% [22]. Under five mortality rate is among the highest in sub-Saharan Africa, with 96 deaths per 1,000 live births [10]. Total fertility rate was estimated at 5.7 and 32.3% of women in union had unmet need for family planning in 2018 [10]. The country recommends a birth interval of 2 to 5 years [23], and the median observed birth interval is 34.1 months [10].

### Study design

This is a quantitative cross-sectional study using secondary data. We carried out analysis of the most recent Benin Demographic and Health Survey (BDHS) from 2017-18.

### Sampling strategy and population of interest

The survey used a two-stage stratified random sampling strategy. The sample is representative at the national level for both urban and rural areas. In the first stage, 555 clusters were selected from the list of enumeration areas established during the fourth General Population and Housing Census conducted in Benin in 2013; by making a systematic sampling with probability proportional to the number of households in the cluster (the size of the clusters is the number of households). Then, a complete household count in each of these clusters provided a list of households from which a target sample of 26 households per cluster was drawn. The details of the survey sampling are given elsewhere [10]. Out of 14,435 households sampled, 99% were surveyed [10]. All women aged 15–49 who were either permanent residents of the sampled households or visitors present in the households on the night before the survey were eligible to be interviewed.

### Study sample size

Our study sample consisted of children aged 6-59 months assessed for stunting at the time of the survey whose mothers were interviewed using the questionnaire for women 15-49 years. Among the 10,292 children, we excluded 139 children who were visitors in the household at the time of survey. The final sample size was 10,153 children.

### Data collection

Questionnaires for women 15-49 years were used to collect information on respondent’s background, contraception at the time of the survey, and birth history. Women were also asked to provide information on the health and nutritional status of their children under five. The information related to food diversity was collected only for children under two years. All children included in the survey were weighed and measured to determine their nutritional status (wasting, underweight, and stunting). Weight was measured with a double-weighing electronic scale (SECA type). Height was measured with a graduated scale (ShorrBoards®). The height of children aged 36-59 months was measured in an upright position. The age of the children was collected from an administrative document or estimated based on special events [10]. In one out of every two households, all women aged 15–49 were measured for weight and height.

### Variables included in the study

#### Outcome variable

Table 1 presents detailed definitions of variables. The outcome is stunting defined as a height for- age z-score lower than 2 standard deviations from the median of the WHO child growth standards [25]. The DHS used data collected on every child’s (height, age, and sex) to calculate the height for- age Z-scores [25] accordingly to the WHO standard growth charts for children.

**Table 1.**
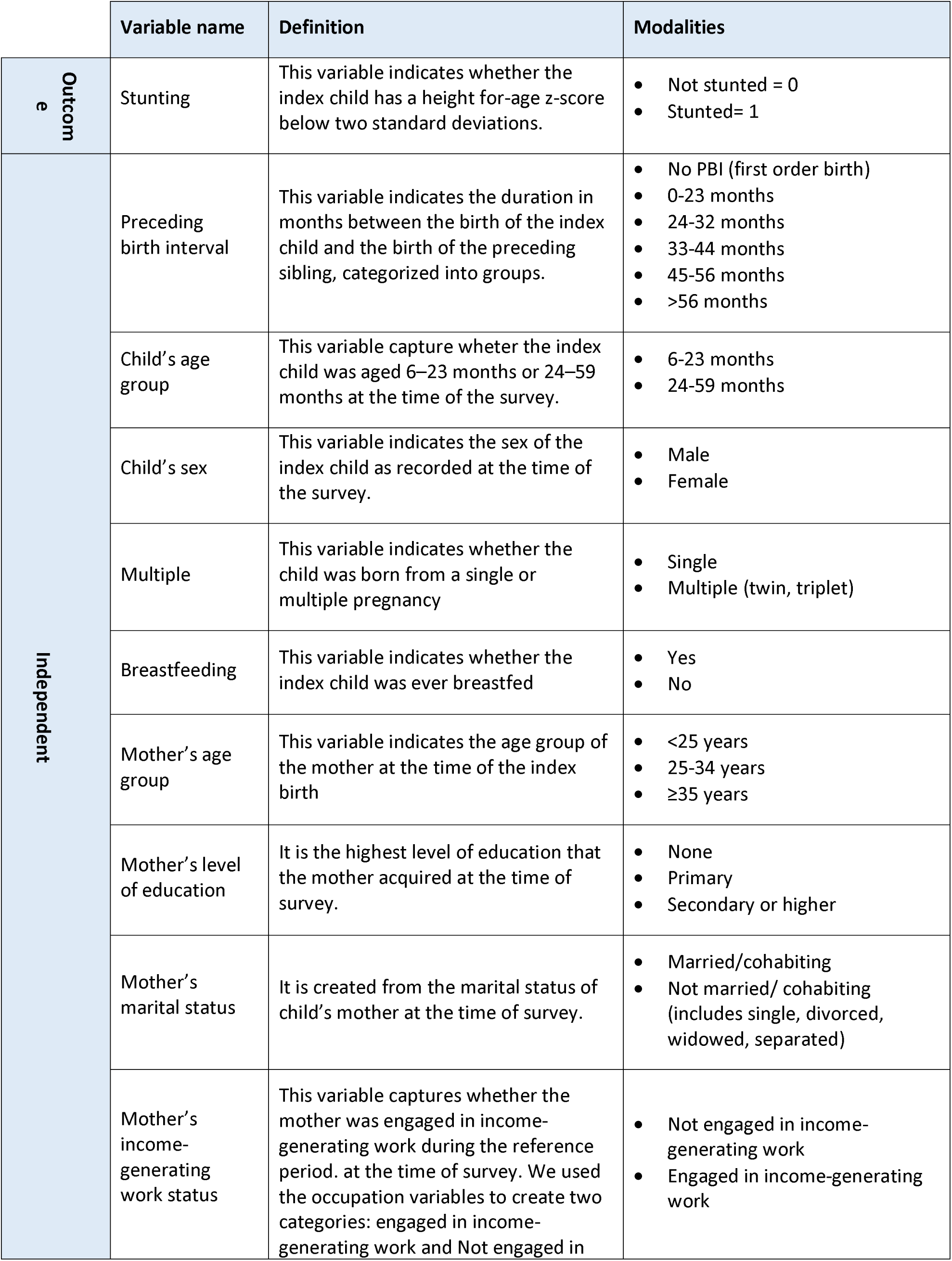
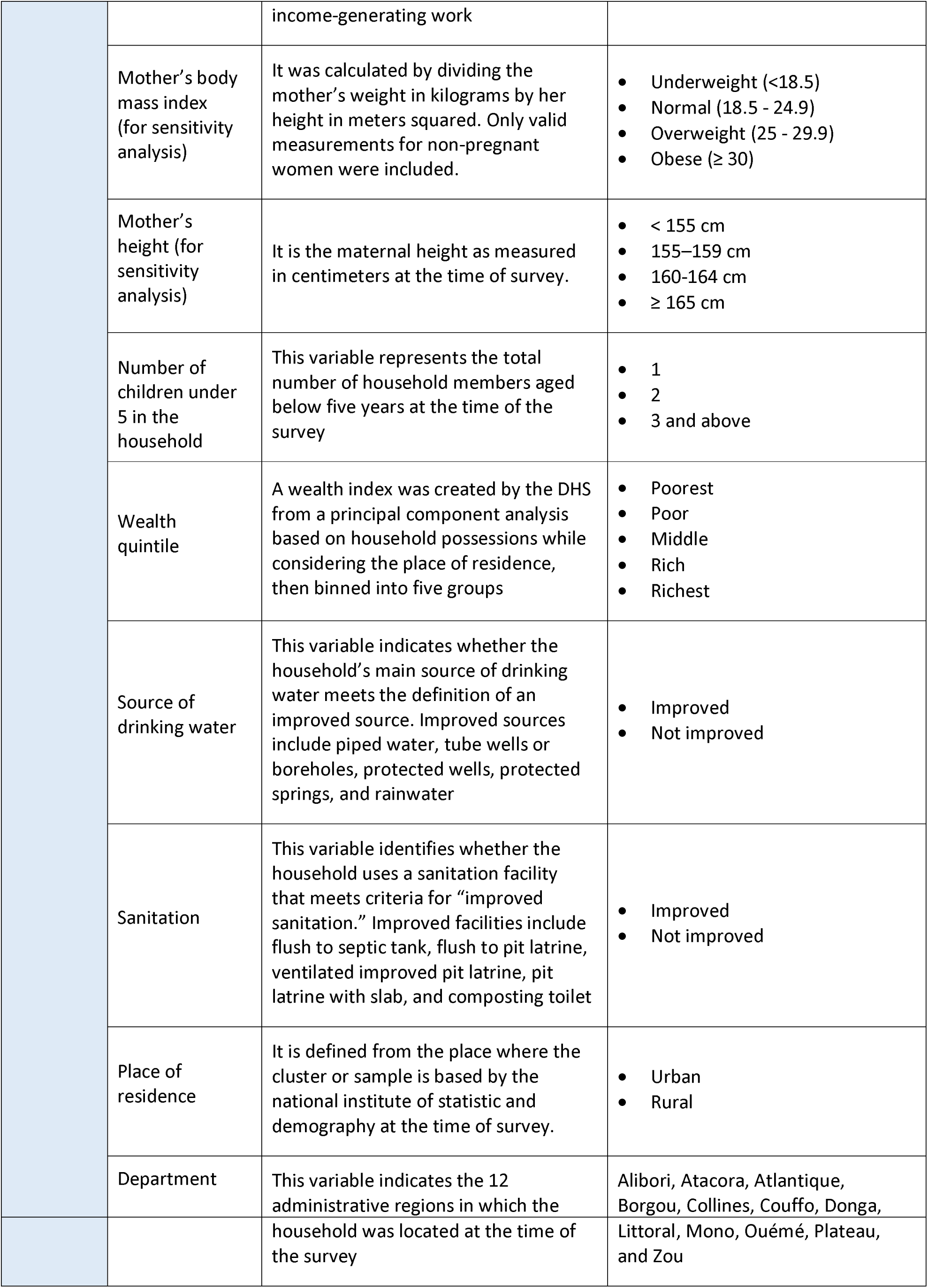
Definitions of variables included in the study.

#### Independent variables

The primary independent variable was PBI, defined as the duration in months between the index child’s birth and the birth of the preceding childbirth of the mother [26]. The interval from previous birth to index pregnancy (conception) was not directly measured. Although we could use reported birth dates and gestational duration to capture it, such approach could introduce additional measurement error due to recall bias and uncertainty in gestational dating. Therefore, the preceding birth-to-birth interval, derived from reported dates of birth, was used. We categorized PBI variable into six groups: (i) less than 24 months; (ii) 24-32 months; (iii) 33-44 months; (iv) 45-56 months; (v) more than 56 months and (vi) no PBI (first born) to capture the different options used in previous research as well as WHO recommendation [14,15,27]. Although the WHO recommends a minimum interpregnancy interval of 24 months, this corresponds approximately to a birth-to-birth interval of 33 months when accounting for the duration of a full-term pregnancy. Therefore, we used 33 months as threshold to align our PBI categorization with this recommendation. Fig 1 below highlight PBI in the birth story of a woman.

**Fig 1.**
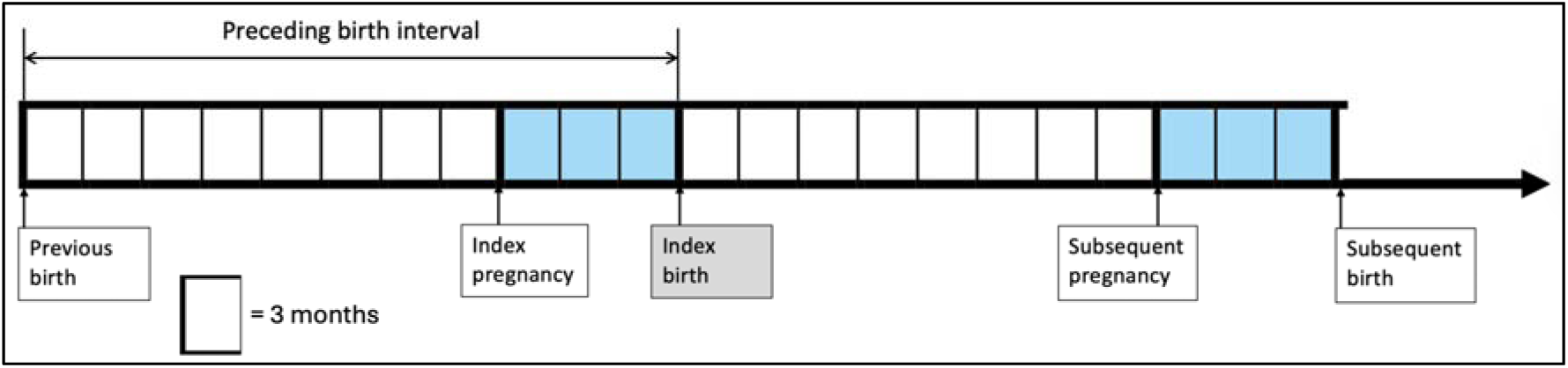
Birth interval description, adapted from WHO [17] and Dewey et al. [16].

We included 14 potential confounders related to child (sex, whether the child was born from a single or multiple pregnancy, birth order, whether the child was ever breastfed), mother (age group at birth of index child, level of education, marital status, employment status), and household-level characteristics (number of children under 5 in the household, source of water, sanitation, wealth index, place of residence and department). Our work was guided by an adapted version of the Vaiada et al conceptual framework highlighting multi-level determinants of stunting [8], and inspired by other literature [4,9,15,27]. As women aged 15–49 were measured for weight and height in one out of every two households, this result in 53.9% missing data for mother body mass index and height. These variables are strongly associated with child stunting [4,8,9], and we therefore conducted a sensitivity analysis by including the variables in the model, which showed no major changes in findings (Supplementary Material 1). We report the final models excluding the mother’s height and body mass index to preserve sample size.

### Data analysis

We first provide a description of the sample according to children’s socio-demographic characteristics, those of their mother and households. Frequencies and percentages with 95% confidence intervals (CI) were summarized. Results were presented in tables and visualized using graphs. Associations between stunting and independent variables were explored with binary logistic regression and reported through unadjusted odds ratios (OR) with 95% confidence intervals and p-value. Independent variables with p-value <0.2 were included in a multi-variable logistic regression. We introduced a multiplicative interaction term between PBI and child age group to evaluate whether the association between PBI and stunting varied by child age group, and compared it against the main-effects-only specification. Model selection was guided by a convergence of statistical criteria (Hosmer–Lemeshow test; Akaike Information Criterion (AIC), Bayesian Information Criterion (BIC) and likelihood-ratio test derived from the corresponding non-survey-weighted models). We further conducted stratified logistic regression analyses separately for children aged 6– 23 months and 24–59 months to explore the consistency of the observed associations across age groups, comparing stratum-specific results descriptively with those from the pooled model. This analysis was justified by the differential vulnerability during the critical window of growth (first 1,000 days) and the cumulative nature of stunting. Variance inflation factors (VIF) were calculated for all independent variables and VIF values <3 indicated acceptable levels of multicollinearity. Each model specification was assessed using Hosmer–Lemeshow goodness-of-fit test. Statistical significance level was set at p-value<0.05. A sensitivity analysis explored including mother’s body mass index and height showed no differences in the distribution of sample characteristics and association (S3, S4 and S5 Tables). In the sensitivity analysis, we excluded children of women who were reported being pregnant at the time of the survey, as pregnancy alters maternal body mass index and could lead to misclassification of this independent variable. Stata software version 15.0 (StataCorp, College Station, TX, USA) was used for data analysis. Considering the sampling procedure, all analyses were performed using STATA "svy" package, except for the models computed to assess the contribution of interaction terms.

### Ethics

The 2017-18 BDHS obtained the ethical approval of the National Ethics Committee for Health Research of Benin [10]. Informed written consent was obtained from all participants prior to the interview, including parental or guardian consent for the measurement of height and weight of children under five years old [10]. All data were collected and processed in a manner that ensured participant privacy and confidentiality [28]. We applied to the DHS-Program and obtained access and permission to use the database on 14 April 2022. The data accessed were entirely anonymous, and we did not have access to any information that could identify individual participants.

### Patient and public involvement

Secondary data were used for this analysis. Therefore, no patient consent was needed.

## Results

### Sample characteristics

We included 10,153 children; with 3,831 (37.6%) in the 6-23 months age group (Table 2). Most children were born after intervals of 24–32 months (24.1%, 95% CI 23.1–25.2) and 33–44 months (22.8%, 21.9–23.8). Short PBI (<24 months) accounted for 11.6% (10.9–12.3), while 21.7% (20.8–22.6) of children were first born (had no PBI). The majority of mothers were between 25 and 34 years old at the time of the birth of the index child (52.7%), had no formal education (65.4%), had a income-generating work (84.1%), and maried or cohabiting with a partner (94.2%) at the time of the survey. Most children lived in a household with two or more children under 5 (75.8%), resided in rural areas (60.2%), had access to water from an improved source (67.8%), but used unimproved sanitation (71.2%).

**Table 2.** Child, maternal, and household characteristics of the study population, BDHS 2017–2018 (N = 10,153)

| Variables | n | % [95 CI] |
| --- | --- | --- |
| **Preceding birth interval** |  |  |
| No preceding birth interval | 2172 | 21.7 (20.8 - 22.6) |
| <24 | 1183 | 11.6 (10.9 - 12.3) |
| 24-32 | 2438 | 24.1 (23.1 - 25.2) |
| 33-44 | 2336 | 22.8 (21.9 - 23.8) |
| 45-56 | 1019 | 10.0 (9.4 - 10.7) |
| >56 | 1005 | 9.8 (9.1 - 10.5) |
| **Child age group (in months)** |  |  |
| 6-23 | 3831 | 37.6 (36.7 - 38.4) |
| 24-59 | 6322 | 62.4 (61.6 - 63.3) |
| **Child sex** |  |  |
| Male | 5166 | 51 (50 - 52.1) |
| Female | 4987 | 49 (47.9 - 50) |
| **Multiplicity** |  |  |
| Singleton | 9736 | 95.7 (95.2 - 96.2) |
| Multiple | 417 | 4.3 (3.8 - 4.8) |
| **Breastfeeding** |  |  |
| Ever breastfed | 9734 | 96.0 (95.4 - 96.5) |
| Never breastfed | 419 | 4.0 (3.5 - 4.6) |
| **Maternal age at birth of index child** |  |  |
| <25 | 2355 | 23.4 (22.3 - 24.6) |
| 25-34 | 5339 | 52.7 (51.4 - 54.1) |
| >=35 | 2459 | 23.8 (22.7 - 24.9) |
| **Maternal level of education** |  |  |
| None | 6640 | 65.4 (63.4 - 67.3) |
| Primary | 1842 | 18.3 (17.1 - 19.6) |
| Secondary or higher | 1671 | 16.3 (15 - 17.7) |
| **Maternal marital status** |  |  |
| Not married/ cohabiting | 572 | 5.8 (5.1 - 6.5) |
| Married or cohabiting | 9581 | 94.2 (93.5 - 94.9) |
| **Mother income-generating work** |  |  |
| Not engaged in income-generating work | 1603 | 15.9 (14.5 - 17.5) |
| Engaged in income-generating work | 8550 | 84.1 (82.5 - 85.5) |
| **Number of children under 5 in the household** |  |  |
| 1 | 2449 | 24.2 (22.9 - 25.5) |
| 2 | 4194 | 41.7 (40 - 43.4) |
| 3+ | 3510 | 34.1 (32.1 - 36.3) |
| **Household wealth index** |  |  |
| Poorest | 2143 | 20.3 (17.9 - 23) |
| Poor | 1998 | 19.8 (18.4 - 21.4) |
| Middle | 2025 | 20.4 (18.9 - 22) |
| Rich | 2030 | 20.6 (18.9 - 22.3) |
| Richest | 1957 | 18.8 (17 - 20.8) |
| **Source of water** |  |  |
| Improved | 6951 | 67.8 (65 - 70.5) |
| Unimproved | 3202 | 32.2 (29.5 - 35) |
| **Sanitation** |  |  |
| Improved | 7252 | 28.8 (26.5 - 31.2) |
| Unimproved | 2901 | 71.2 (68.8 - 73.5) |
| **Place of residence** |  |  |
| Urban | 4173 | 39.8 (37.1 - 42.5) |
| Rural | 5980 | 60.2 (57.5 - 62.9) |
| **Departement** |  |  |
| Alibori | 1198 | 13.2 (11.3 - 15.5) |
| Atacora | 989 | 8.9 (7.6 - 10.4) |
| Atlantique | 968 | 11.5 (10 - 13.1) |
| Borgou | 1243 | 12.6 (11.1 - 14.3) |
| Collines | 811 | 6.5 (5.6 - 7.5) |
| Couffo | 654 | 6.5 (5.8 - 7.4) |
| Donga | 738 | 6.5 (5.6 - 7.6) |
| Littoral | 687 | 4.7 (3.8 - 5.7) |
| Mono | 514 | 4.6 (4 - 5.2) |
| Ouémé | 805 | 9.2 (8.2 - 10.4) |
| Plateau | 604 | 6.1 (5 - 7.4) |
| Zou | 942 | 9.6 (8.4 - 10.9) |

**Table 3.** Association between PBI and stunting in bivariate and multivariable logistic regression.

| Variables | Unadjusted |  | Adjusted without interaction (Model A)<sup>a</sup> |  | Adjusted with interaction (Model B)<sup>b</sup> |  |
| --- | --- | --- | --- | --- | --- | --- |
|  | OR [95 CI] | p-value | OR [95 CI] | p-value | OR [95 CI] | p-value |
| **PBI in months (Ref: <24)** |  |  |  |  |  |  |
| 24-32 | 0.90 (0.78 - 1.05) | 0.193 | 0.91 (0.77 - 1.06) | 0.218 | 0.89 (0.69 - 1.16) | 0.389 |
| 33-44 | 0.93 (0.8 - 1.08) | 0.350 | 0.95 (0.81 - 1.11) | 0.509 | 0.88 (0.67 - 1.17) | 0.385 |
| 45-56 | 0.59 (0.49 - 0.72) | <0.001 | 0.63 (0.51 - 0.78) | <0.001 | 0.69 (0.49 - 0.96) | 0.029 |
| >56 | 0.56 (0.46 - 0.68) | <0.001 | 0.67 (0.54 - 0.82) | <0.001 | 0.78 (0.56 - 1.1) | 0.159 |
| No PBI | 0.78 (0.66 - 0.92) | 0.004 | 0.85 (0.7 - 1.03) | 0.103 | 0.96 (0.7 - 1.33) | 0.825 |
| **Child age group in months (Ref: 6-23)** | 1.41 (1.29 - 1.55) | <0.001 | 1.47 (1.33 - 1.62) | <0.001 | 1.53 (1.16 - 2) | 0.002 |
| **PBI # Child age group** |  |  |  |  |  |  |
| No PBI # 2. 24-59 |  |  |  |  | 0.83 (0.57 - 1.21) | 0.340 |
| 24-32 #2. 24-59 |  |  |  |  | 1.03 (0.73 - 1.43) | 0.884 |
| 33-44 #2. 24-59 |  |  |  |  | 1.12 (0.79 - 1.58) | 0.526 |
| 45-56 #2. 24-59 |  |  |  |  | 0.89 (0.59 - 1.33) | 0.563 |
| 57-243 #2. 24-59 |  |  |  |  | 0.79 (0.51 - 1.24) | 0.304 |
| **Child sex (Ref: Male)** | 0.79 (0.72 - 0.86) | <0.001 | 0.77 (0.71 - 0.84) | <0.001 | 0.77 (0.71 - 0.84) | <0.001 |
| **Multiple (Ref: Single)** | 2.38 (1.87 - 3.04) | <0.001 | 2.63 (2.05 - 3.38) | <0.001 | 2.62 (2.04 - 3.37) | <0.001 |
| **Breastfeeding (Ref: ever breastfed)** | 0.79 (0.64 - 0.98) | 0.031 | 0.83 (0.67 - 1.03) | 0.088 | 0.83 (0.67 - 1.04) | 0.101 |
| **Maternal age at birth of index child (Ref: <25)** |  |  |  |  |  |  |
| 25-34 | 0.93 (0.82 - 1.04) | 0.193 | 0.85 (0.73 - 0.98) | 0.029 | 0.85 (0.73 - 0.99) | 0.034 |
| >=35 | 0.87 (0.75 - 1) | 0.051 | 0.75 (0.62 - 0.9) | 0.002 | 0.75 (0.62 - 0.9) | 0.003 |
| **Maternal level of education (Ref: None)** |  |  |  |  |  |  |
| Primary | 0.77 (0.68 - 0.88) | <0.001 | 0.95 (0.82 - 1.09) | 0.450 | 0.95 (0.82 - 1.09) | 0.461 |
| secondary or higher | 0.51 (0.44 - 0.59) | <0.001 | 0.73 (0.62 - 0.86) | <0.001 | 0.73 (0.62 - 0.86) | <0.001 |
| **Maternal marital status (Ref: not married/cohabiting)** | 0.94 (0.77 - 1.13) | 0.488 |  |  |  |  |
| **Mother working status (Ref: no income generating work)** | 0.97 (0.86 - 1.1) | 0.605 |  |  |  |  |
| **Number of children under 5 in the household (Ref: 1)** |  |  |  |  |  |  |
| 2 | 1.23 (1.1 - 1.39) | 0.001 | 1.03 (0.9 - 1.17) | 0.707 | 1.06 (0.92 - 1.21) | 0.425 |
| +3 | 1.56 (1.38 - 1.77) | <0.001 | 1.13 (0.98 - 1.3) | 0.086 | 1.16 (1.01 - 1.34) | 0.042 |
| **Household wealth index (Ref: Poorest)** |  |  |  |  |  |  |
| Poor | 0.86 (0.74 - 1) | 0.047 | 0.88 (0.76 - 1.02) | 0.085 | 0.88 (0.76 - 1.02) | 0.088 |
| Middle | 0.68 (0.58 - 0.79) | <0.001 | 0.71 (0.6 - 0.84) | <0.001 | 0.71 (0.6 - 0.84) | <0.001 |
| Rich | 0.53 (0.45 - 0.62) | <0.001 | 0.58 (0.48 - 0.69) | <0.001 | 0.58 (0.49 - 0.69) | <0.001 |
| Richest | 0.31 (0.26 - 0.38) | <0.001 | 0.38 (0.29 - 0.49) | <0.001 | 0.38 (0.29 - 0.49) | <0.001 |
|  | OR [95 CI] | p-value | OR [95 CI] | p-value | OR [95 CI] | p-value |
| Improved source of water (Ref: Unimproved) | 0.76 (0.68 - 0.86) | <0.001 | 0.97 (0.86 - 1.09) | 0.579 | 0.97 (0.86 - 1.09) | 0.575 |
| Improved sanitation (Ref: Unimproved) | 0.55 (0.49 - 0.63) | <0.001 | 0.89 (0.76 - 1.05) | 0.163 | 0.89 (0.76 - 1.05) | 0.160 |
| Place of residence (Ref: Urban) | 1.44 (1.26 - 1.64) | <0.001 | 1.03 (0.9 - 1.17) | 0.679 | 1.03 (0.9 - 1.17) | 0.686 |
| Departement (Ref: Littoral) |  |  |  |  |  |  |
| Alibori | 2.68 (1.9 - 3.77) | <0.001 | 1 (0.69 - 1.43) | 0.985 | 1 (0.7 - 1.43) | 0.996 |
| Atacora | 2.44 (1.72 - 3.45) | <0.001 | 0.91 (0.64 - 1.31) | 0.626 | 0.92 (0.64 - 1.31) | 0.632 |
| Atlantique | 1.89 (1.34 - 2.67) | <0.001 | 1.08 (0.77 - 1.53) | 0.644 | 1.09 (0.77 - 1.53) | 0.633 |
| Borgou | 2.29 (1.61 - 3.25) | <0.001 | 0.95 (0.68 - 1.34) | 0.788 | 0.96 (0.68 - 1.35) | 0.805 |
| Collines | 1.37 (0.95 - 1.98) | 0.093 | 0.63 (0.44 - 0.92) | 0.015 | 0.64 (0.44 - 0.92) | 0.017 |
| Couffo | 2.56 (1.77 - 3.69) | <0.001 | 1.21 (0.84 - 1.74) | 0.310 | 1.21 (0.84 - 1.74) | 0.314 |
| Donga | 1.54 (1.03 - 2.29) | 0.036 | 0.71 (0.49 - 1.05) | 0.083 | 0.71 (0.49 - 1.04) | 0.081 |
| Mono | 1.76 (1.18 - 2.62) | 0.006 | 0.85 (0.57 - 1.26) | 0.423 | 0.86 (0.58 - 1.27) | 0.439 |
| Ouémé | 1.73 (1.21 - 2.47) | 0.003 | 1.08 (0.76 - 1.53) | 0.678 | 1.08 (0.76 - 1.53) | 0.681 |
| Plateau | 2.46 (1.69 - 3.58) | <0.001 | 1.14 (0.78 - 1.67) | 0.496 | 1.14 (0.78 - 1.67) | 0.498 |
| Zou | 2.36 (1.67 - 3.35) | <0.001 | 1.23 (0.88 - 1.72) | 0.230 | 1.23 (0.88 - 1.72) | 0.227 |

<sup>a</sup>Model A: Goodness-of-fit test (p-value=0.2790); AIC(model without svy)=12333.53; BIC(model without svy)=12579.2
<sup>b</sup>Model B: Goodness-of-fit test (p-value=0.1832) ; AIC(model without svy)=12337.82; BIC(model without svy)=12619.62

### Prevalence of stunting by children’s age and PBI

Fig 2 presents the percentage of children who were stunted, along with their 95% confidence intervals, disaggregated by PBI and age group. The overall prevalence of stunting was 33.5% (95% CI: 32.1-34.9%), with higher percentages observed among children aged 24-59 months (36.3%, 95% CI: 34.7-38.1%) compared to younger children (28.8%, 95% CI: 27.1-30.5%), p<0.001. Among children aged 6–23 months, no significant difference in stunting prevalence was observed across PBI (p-value = 0.073). In the subgroup aged 24–59 months, stunting was lower among those with a PBI of 45–56 months (28.1%; 95% CI: 24.3–32.1) and more than 56 months (26.9%; 95% CI: 23.1–31.1). Stunting was higher among children with PBI of less than 24 months (41.2%; 95% CI: 37.4–45.1), 24–32 months (39.1%; 95% CI: 36.2–42.1), and 33–44 months (40.5%; 95% CI: 37.6–43.5). These differences across PBI were statistically significant (p < 0.001).

**Fig 2.**
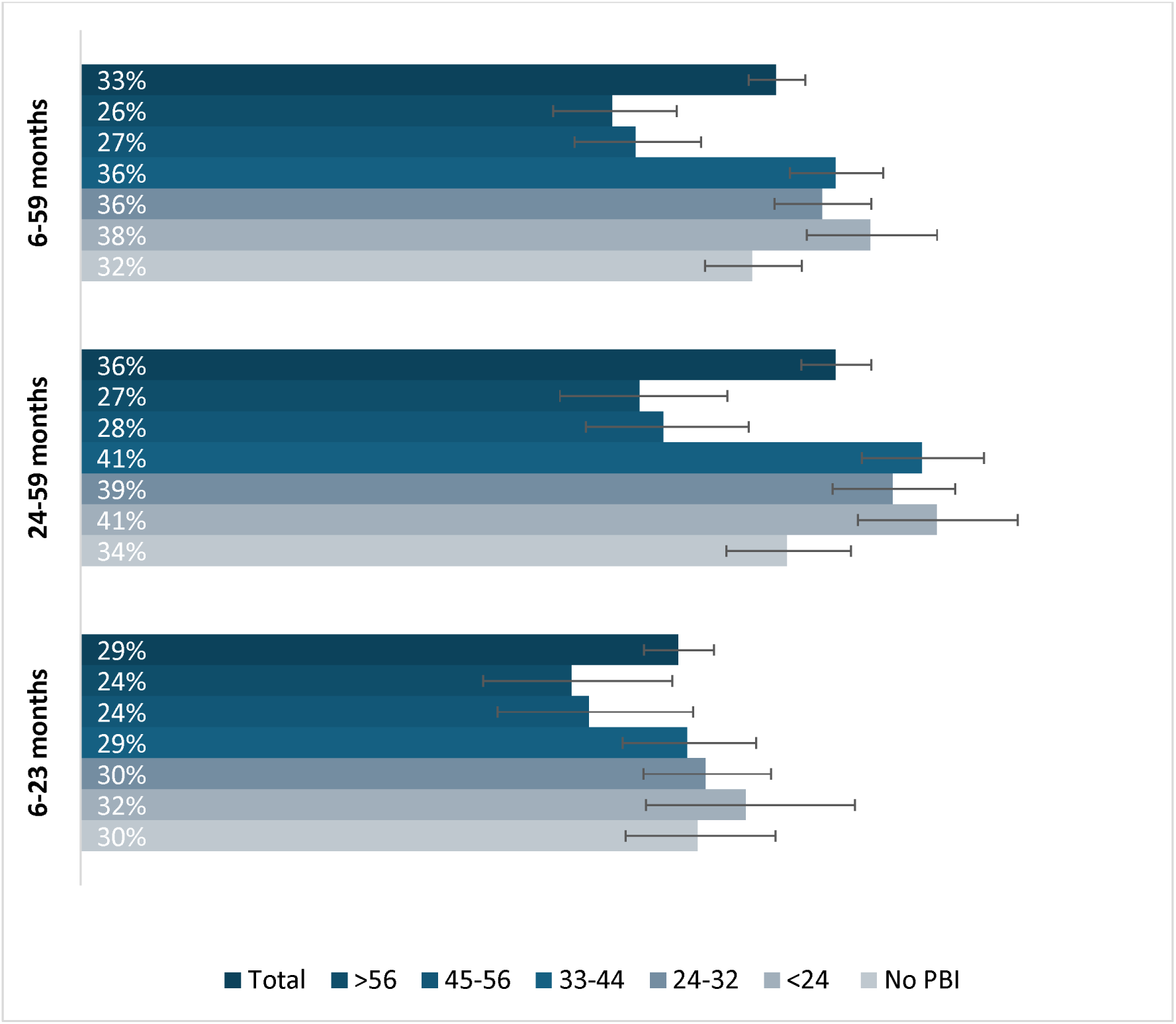
Prevalence of stunting among children and 95% confidence intervals, by previous birth interval category and children’s age group.

### Asociation between PBI and stunting

To determine the most appropriate model specification, a main-effects model (Model A) was compared against an interaction model incorporating a multiplicative term between PBI and child age group (Model B). Model B yielded higher AIC (12337.82 vs. 12333.53) and higher BIC (12619.62 vs. 12579.20) relative to Model A, and a likelihood-ratio test confirmed that the interaction block contributed no statistically significant explanatory power beyond the main effects (p = 0.336). In the survey-adjusted estimates, none of the five PBI × child age group interaction terms reached statistical significance (all p > 0.30), and all corresponding 95% confidence intervals included the null, providing no evidence of effect modification by child age group. On grounds of statistical parsimony, model fit, and epidemiological interpretability, Model A was retained as the final model.

Compared with children born after a PBI of fewer than 24 months, those born after an interval of 45 to 56 months had significantly lower odds of stunting (AOR: 0.63; 95% CI: 0.51–0.78; p < 0.001), as did those born after an interval exceeding 56 months (AOR: 0.67; 95% CI: 0.54–0.82; p < 0.001). By contrast, intervals of 24 to 32 months (AOR: 0.91; 95% CI: 0.77–1.06; p = 0.218) and 33 to 44 months (AOR: 0.95; 95% CI: 0.81–1.11; p = 0.509) were not significantly associated with the odds of stunting relative to the reference category. Similarly, first-born children, for whom no preceding birth interval exists, did not have significantly different odds of stunting compared with children born after an interval of fewer than 24 months (AOR: 0.85; 95% CI: 0.70–1.03; p = 0.103). The results of the sensitivity analysis restricted to children with complete anthropometric data and non-pregnant mothers at the time of survey (n = 4,680) were consistent with those of the main analysis. The protective associations of a PBI of 45–56 months (AOR: 0.54; 95% CI: 0.40–0.72; p < 0.001) and of more than 56 months (AOR: 0.64; 95% CI: 0.47– 0.87; p = 0.004) remained statistically significant (S2 Table). The direction and magnitude of estimates across all PBI categories were preserved, with the additional emergence of a statistically significant protective association for the 24–32 month interval category (AOR: 0.79; 95% CI: 0.62–0.99; p = 0.045).

Stratified analyses conducted separately for children aged 6–23 months and 24–59 months yielded results broadly consistent with those from the pooled model (Fig 3). Shorter intervals of 24–32 months and 33-44 months were not significantly associated with stunting in either child age group. Among children aged 24–59 months, birth intervals of 45–56 months and exceeding 56 months were associated with lower odds of stunting (AOR: 0.63; 95% CI: 0.49-0.82; p<0.001 and AOR: 0.65; 95% CI: 0.49-0.85; p=0.002, respectively). Among children aged 6–23 months, birth intervals of 45–56 months were associated with lower odds of stunting (AOR: 0.62; 95% CI: 0.44–0.88; p<0.007).

**Fig 3.**
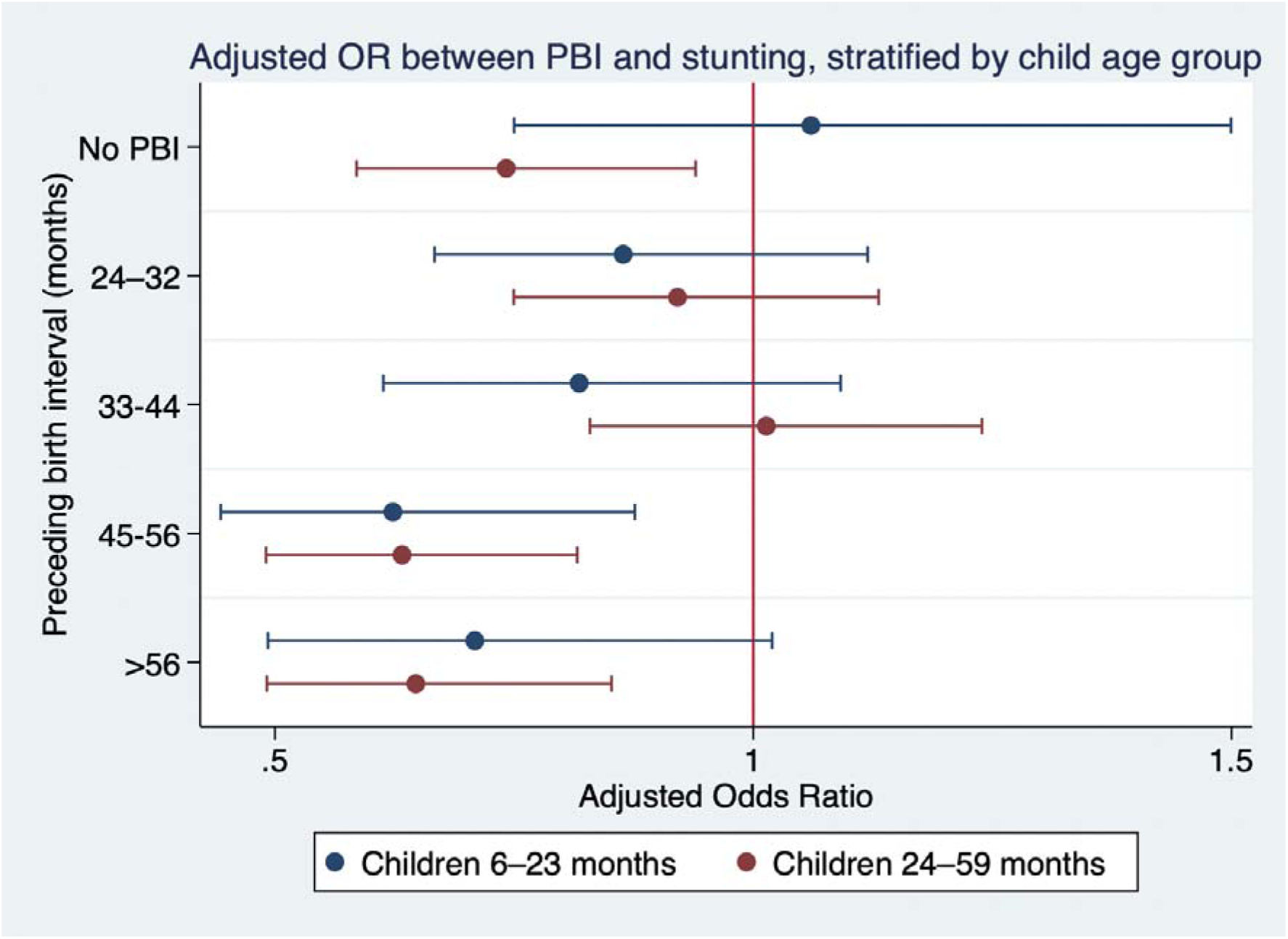
Adjusted association between preceding birth interval and stunting, stratified by child age group.

The sensitivity analysis results were consistent with those from the main analysis. Among children aged 6–23 months (n=1,890), the protective association of birth intervals of 45–56 months remained statistically significant (AOR: 0.56; 95% CI: 0.34–0.94; p = 0.027), and the direction of estimates across all PBI categories were also preserved. The threshold pattern was similarly maintained among children aged 24–59 months, with intervals of 45–56 months (AOR: 0.55; 95% CI: 0.37–0.81; p = 0.002) and exceeding 56 months (AOR: 0.58; 95% CI: 0.40–0.85; p = 0.006) remaining significantly associated with lower odds of stunting. Stratum-specific estimates are presented in supplementary materials (S2, S3 and S4 Tables).

## Discussion

This study examined the association between PBI and stunting among children aged 6–59 months in Benin, using data from the most recent BDHS. We found an absence of a protective effect for the 24- 32 and 33–44 months intervals, even though they exceed the WHO’s minimum recommendation of 24 months following a live birth. After adjusting for potential confounders, children born after an interval of more than 45 months were significantly less likely to be stunted, and the longer the interval, the stronger the association. This finding suggests the potential existence of a longer, context-specific threshold in Benin—around 45 months—beyond which the protective mechanisms against stunting become effective.

Studies conducted in other sub-Saharan African countries reported varying thresholds. In Ethiopia and Ghana, a protective effect was observed from 36 months [15,29]. A meta-analysis drawing on studies from low-and middle-income countries, mainly in Africa and Asia, found an optimum interval in the range of 36–48 months, to be associated with a lower risk of stunting and underweight for children under five years [14]. Such cross-country variability may reflect both methodological heterogeneity across studies — including differences in PBI categorisation, covariate adjustment, and child age groups considered — and contextual factors. The latter may encompass differences in food security, access to healthcare, cultural infant feeding practices, maternal occupation, and baseline maternal nutritional status.

The “maternal depletion syndrome” concept remains relevant in interpreting our findings. Successive pregnancies and breastfeeding episodes deplete maternal stores of essential micronutrients—especially iron, folate, calcium, zinc, and vitamins A and B12 [16]. Replenishment of iron stores typically requires 18–24 months after delivery [16,17], but this period may be considerably extended in contexts of poor dietary intake and continuous physiological demands (e.g., prolonged breastfeeding, strenuous labor). The national prevalence of food insecurity in Benin was estimated at 25.5% in 2022 and nearly 82.6% of households unable to afford a healthy diet in 2021 [30,31]. In the Beninese context where women perform 60-80 % of agricultural work and 44 % of the labor required to feed their families, energy requirements may be elevated, which could further delay recovery [31]. An interval of 45 months or more may provide a sufficient temporal window for mothers to restore nutritional reserves, improve body mass index as our results show, and enter the next pregnancy in optimal physiological condition, favouring fetal development and breast milk quality [32,14]. The absence of a significant association between first-born status and stunting may be explained by primiparity-related risks, including young maternal age and limited experience with pregnancy, infant feeding, and childcare practices, which may partially offset the advantage conferred by the absence of prior maternal nutritional depletion [16,32].

### Strengths and limitations

The use of nationally representative BDHS 2017–2018 data, collected through internationally standardized and quality-assured methods, ensures data reliability. The large sample size of 10,153 children aged 6–59 months from all departements of Benin allows for national representativeness. Adjustments were made for a set of potential confounders at the child, maternal (age, education, BMI, employment status), household levels (socioeconomic level, composition, number of children under five). Stratifying analyses by age group (6–23 vs. 24–59 months) enabled detection of age-specific patterns.

Despite these strengths, some limitations must be acknowledged. Stunting is a multifactorial outcome resulting from the complex interplay of genetic, environmental, and socio-economic factors operating at the individual, household, and community levels [4,8]. While this study focuses on PBI, other important determinants—such as maternal nutritional status, recurrent infections, infant feeding practices, household food security, and caregiving environments—also contribute to child growth. In addition, genetic predispositions and broader environmental conditions, including access to health services, may influence linear growth trajectories. Given this complexity, the observed associations should not be interpreted as causal in isolation. The study may also be subject to residual confounding from unmeasured or imperfectly measured factors at both individual and family levels. While we were able to somewhat account for infant feeding through variables whether the child was ever breastfed and proxy indicators including maternal education and household wealth [33,34], more complex dimensions of feeding practices, such as breastfeeding exclusivity, timing of complementary feeding, and dietary diversity, were not directly controlled for. This indirect adjustment is unlikely to fully account for the variability in feeding behaviors, and may therefore result in incomplete control of their effects, potentially biasing the estimated association between PBI and stunting. In addition, the data are from 2017–2018; although relatively recent, subsequent changes in Benin’s health, nutrition, and socioeconomic landscape—such as expanded social protection programs, improved access to family planning, or the effects of shocks like the COVID-19 pandemic and climate change on food security—could influence the present-day relationship between birth spacing and stunting.

### Implications for research, policy, and practice in Benin

The absence of a protective effect for birth intervals up to 44 months following a previous birth highlights the need of context-specific, data-driven recommendations on optimal birth spacing for maternal and child health. Public health messages on birth spacing should therefore be reframed to emphasize not only the minimum threshold but also the potential added benefits of longer intervals. The findings also underscore the need to strengthen family planning programs in Benin. With 32.3% of women in union experiencing unmet need for family planning in 2018, there remains substantial room for improvement.

## Conclusion

This study showed that children born with longer PBI (≥45 months) have a significantly reduced risk of stunting among children aged 6–59 months in Benin. The absence of a protective effect for the 33–44-month interval—despite exceeding the WHO’s minimum recommendation—highlights the need for context-specific adaptation of global guidelines. It also underscores the need for complementary analyses to determine a more precise spacing threshold suited to Benin’s specific context.

## Acknowledgments

We acknowledge the women who participated in the BDHS 2017-18 for their time and contributions.

## Data availability statement

Data are available in a public, open access repository. Available: https://dhsprogram.com/data/.

## Authors contributions

Conceptualization: Mahugnon Christian Urlyss Agossou, Lenka Beňová Data curation: Mahugnon Christian Urlyss Agossou

Formal analysis: Mahugnon Christian Urlyss Agossou

Methodology: Mahugnon Christian Urlyss Agossou, Giulia Scarpa, Kerry Wong, Lenka Beňová Supervision: Lenka Beňová, Kerry Wong, Giulia Scarpa, Christelle Boyi Hounsou, Jean-Paul Dossou Visualization: Mahugnon Christian Urlyss Agossou

Writing – original draft: Mahugnon Christian Urlyss Agossou

Writing – review & editing: Mahugnon Christian Urlyss Agossou, Giulia Scarpa, Lenka Benova, Diana Sagastume, Christelle Boyi Hounsou, Gottfried Agballa, Jean-Paul Dossou, Kerry Wong

## Supporting information

S1 Table. Sample characteristics of the study population used in the sensitivity analyses (n = 4,680), 2017-18 BDHS.

(DOCX)

S2 Table. Survey-adjusted logistic regression models for the association between preceding birth interval and stunting: main-effects model (Model 1A) versus interaction model (Model 2A) among children aged 6–59 months, sensitivity analysis sample (n = 4,680), 2017-18 Benin DHS.

(DOCX)

S3 Table. Survey-adjusted logistic regression models for the association between preceding birth interval and stunting among children aged 6–23 months: core analysis sample (Model M23, n = 3,831) and sensitivity analysis sample (Model M23A, n = 1,890), Benin DHS.

(DOCX)

S4 Table. Survey-adjusted logistic regression models for the association between preceding birth interval and stunting among children aged 24–59 months: core analysis sample (Model M59, n = 6,322) and sensitivity analysis sample (Model M59A, n = 2,790), Benin DHS.

(DOCX)

