## Supplemental table 1 to table 4 for "Is a 24-Month Birth Interval Enough? Evidence from a cross-sectional study using the Benin Demographic and Health Survey"

**S1 Table. Sample characteristics of the study population used in the sensitivity analyses (n = 4,680), 2017-18 BDHS.**

| **Variables** | **n** | **% (95% CI)** |
| --- | --- | --- |
| **Preceding birth interval (months)** | | |
| < 24 months | 551 | 11.8 (10.6–12.7) |
| 24–32 months | 1,087 | 23.5 (22.1–24.9) |
| 33–44 months | 1,111 | 23.5 (22.1–25.0) |
| 45–56 months | 476 | 10.2 (9.3–11.3) |
| > 56 months | 481 | 10.1 (9.1–11.1) |
| No preceding birth (first-born) | 974 | 21.1 (19.8–22.5) |
| **Child age group** | | |
| 6–23 months | 1,890 | 40.4 (39.1–41.7) |
| 24–59 months | 2,790 | 59.6 (58.3–60.9) |
| **Child sex** | | |
| Male | 2,387 | 51.4 (49.9–52.8) |
| Female | 2,293 | 48.6 (47.2–50.1) |
| **Birth type** | | |
| Single | 4,467 | 95.5 (94.4–96.1) |
| Multiple | 213 | 4.5 (3.9–5.6) |
| **Ever breastfed** | | |
| Yes | 4,484 | 95.9 (95.1–96.5) |
| No | 196 | 4.1 (3.5–4.9) |
| **Maternal age at birth of index child (years)** | | |
| 20–24 | 1,050 | 22.7 (21.2–24.3) |
| 25–34 | 2,437 | 52.2 (50.2–54.2) |
| ≥ 35 | 1,193 | 25.1 (23.5–26.8) |
| **Maternal level of education** | | |
| None (Ref) | 3,075 | 65.8 (63.5–68.0) |
| Primary | 814 | 17.4 (16.0–19.2) |
| Secondary or above | 791 | 16.7 (15.1–18.4) |
| **Maternal marital status** | | |
| Not married / cohabiting | 307 | 6.5 (5.7–7.5) |
| Married / cohabiting | 4,373 | 93.5 (92.5–94.3) |
| **Mother income-generating working status** | | |
| Not engaged in income-generating work | 707 | 15.1 (13.3–16.8) |
| Engaged in income-generating work | 3,973 | 84.9 (83.2–86.7) |
| **Maternal BMI (kg/m²)** | | |
| Underweight | 407 | 8.6 (7.5–9.8) |
| Normal | 3,076 | 65.9 (63.9–67.8) |
| Overweight | 814 | 17.4 (15.9–19.1) |
| Obese | 383 | 8.1 (6.9–9.3) |
| **Maternal height** | | |
| < 155 cm | 1,166 | 24.9 (23.2–26.7) |
| 155–159 cm | 1,610 | 34.5 (32.6–36.5) |
| 160–164 cm | 1,196 | 25.6 (23.7–26.9) |
| ≥ 165 cm | 708 | 15.1 (13.9–16.7) |
| **Number of children under 5 in household** | | |
| 1 | 1,069 | 22.8 (21.2–24.4) |
| 2 | 1,913 | 40.9 (39.2–43.7) |
| ≥ 3 | 1,698 | 36.3 (33.2–38.6) |
| **Household wealth index** | | |
| Poorest | 925 | 18.6 (16.3–21.1) |
| Poorer | 876 | 18.9 (17.1–20.9) |
| Middle | 963 | 21.3 (19.3–23.5) |
| Richer | 967 | 21.5 (19.5–23.7) |
| Richest | 949 | 19.7 (17.6–21.9) |
| **Source of drinking water** | | |
| Unimproved | 1,406 | 30.5 (27.6–33.7) |
| Improved water source | 3,274 | 69.5 (66.3–72.4) |
| **Sanitation facility** | | |
| Unimproved sanitation | 3,258 | 69.6 (66.7–72.1) |
| Improved sanitation | 1,422 | 30.4 (27.9–33.3) |
| **Place of residence** | | |
| Urban | 1,901 | 38.8 (35.9–41.6) |
| Rural | 2,779 | 61.2 (58.4–64.1) |
| **Département** | | |
| Alibori | 554 | 13.4 (11.4–15.6) |
| Atacora | 453 | 9.1 (7.5–10.8) |
| Atlantique | 437 | 11.1 (9.6–12.9) |
| Borgou | 541 | 11.9 (10.4–13.6) |
| Collines | 372 | 6.5 (5.5–7.6) |
| Couffo | 313 | 6.7 (5.8–7.8) |
| Donga | 320 | 6.2 (5.1–7.4) |
| Littoral | 355 | 5.0 (4.2–6.0) |
| Mono | 244 | 4.8 (4.1–5.6) |
| Ouémé | 372 | 9.3 (8.1–10.6) |
| Plateau | 242 | 5.4 (4.3–6.8) |
| Zou | 477 | 10.6 (9.1–12.3) |
| **Stunting status** | | |
| No | 3,103 | 66.7 (64.9–68.4) |
| Yes | 1,577 | 33.3 (31.6–35.1) |

Note: The sensitivity analysis sample (n = 4,680) is a subset of the main analytic sample (n = 10,153) restricted to children with complete data on maternal BMI, height, and without currently pregnant mothers.

Abbreviations: CI = Confidence interval; BMI = Body mass index; SD = Standard deviation; DHS = Demographic and Health Survey.

**S2 Table. Survey-adjusted logistic regression models for the association between preceding birth interval and stunting: main-effects model (Model 1A) versus interaction model (Model 2A) among children aged 6–59 months, sensitivity analysis sample (n = 4,680), 2017-18 Benin DHS.**

| **Variables** | **Model 1A (n = 4,680)** | | **Model 2A — With interaction (n = 4,680)** | |
| --- | --- | --- | --- | --- |
|  | **AOR (95% CI)** | **p-value** | **AOR (95% CI)** | **p-value** |
| **Preceding birth interval (months) (Ref: < 24 months)** | | | | |
| *< 24 months (Ref)* | — |  | — |  |
| 24–32 months | 0.79 (0.62–0.99) | 0.045 | 0.73 (0.47–1.13) | 0.157 |
| 33–44 months | 0.92 (0.73–1.17) | 0.502 | 0.85 (0.55–1.32) | 0.469 |
| 45–56 months | 0.54 (0.40–0.72) | <0.001 | 0.64 (0.39–1.05) | 0.076 |
| > 56 months | 0.64 (0.47–0.87) | 0.004 | 0.91 (0.53–1.55) | 0.721 |
| No preceding birth (first-born) | 0.81 (0.62–1.04) | 0.102 | 1.04 (0.67–1.63) | 0.857 |
| **Child age group (Ref: 6–23 months)** | | | | |
| *6–23 months (Ref) (Ref)* | — |  | — |  |
| 24–59 months | 1.57 (1.36–1.83) | <0.001 | 1.72 (1.12–2.63) | 0.012 |
| **PBI × Child age group interaction (Ref: < 24 months # 6–23 months)** | | | | |
| 24–32 months # 24–59 months | — | — | 1.13 (0.68–1.89) | 0.634 |
| 33–44 months # 24–59 months | — | — | 1.15 (0.69–1.91) | 0.593 |
| 45–56 months # 24–59 months | — | — | 0.78 (0.41–1.49) | 0.456 |
| > 56 months # 24–59 months | — | — | 0.59 (0.30–1.17) | 0.132 |
| No PBI # 24–59 months | — | — | 0.68 (0.39–1.17) | 0.161 |
| **Child sex (Ref: Male)** | | | | |
| Female | 0.79 (0.70–0.90) | 0.001 | 0.79 (0.69–0.90) | 0.001 |
| **Birth type (Ref: Single)** | | | | |
| Multiple | 3.16 (2.22–4.49) | <0.001 | 3.13 (2.19–4.47) | <0.001 |
| **Ever breastfed (Ref: No)** | | | | |
| Yes | 0.85 (0.61–1.18) | 0.335 | 0.85 (0.62–1.18) | 0.343 |
| **Maternal age at birth (years) (Ref: 20–24)** | | | | |
| 25–34 | 0.80 (0.64–1.00) | 0.050 | 0.80 (0.64–1.00) | 0.054 |
| ≥ 35 | 0.76 (0.58–0.99) | 0.040 | 0.76 (0.58–0.99) | 0.041 |
| **Maternal level of education (Ref: None)** | | | | |
| Primary | 0.84 (0.69–1.04) | 0.104 | 0.85 (0.69–1.04) | 0.117 |
| Secondary or above | 0.75 (0.59–0.95) | 0.017 | 0.75 (0.59–0.95) | 0.017 |
| **Maternal BMI (Ref: Thin < 18.5 kg/m²)** | | | | |
| Normal (18.5–24.9) | 0.77 (0.60–0.99) | 0.045 | 0.77 (0.60–1.00) | 0.046 |
| Overweight (25.0–29.9) | 0.58 (0.42–0.79) | 0.001 | 0.58 (0.43–0.80) | 0.001 |
| Obese (≥ 30.0) | 0.43 (0.28–0.64) | <0.001 | 0.43 (0.29–0.64) | <0.001 |
| **Maternal height (Ref: < 155 cm)** | | | | |
| 155–159 cm | 0.69 (0.56–0.84) | <0.001 | 0.69 (0.57–0.84) | <0.001 |
| 160–164 cm | 0.35 (0.28–0.43) | <0.001 | 0.34 (0.28–0.43) | <0.001 |
| ≥ 165 cm | 0.36 (0.27–0.46) | <0.001 | 0.36 (0.27–0.47) | <0.001 |
| **Number of children under 5 in household (Ref: 1)** | | | | |
| 2 | 1.14 (0.93–1.40) | 0.197 | 1.22 (0.99–1.51) | 0.063 |
| ≥ 3 | 1.21 (0.98–1.49) | 0.073 | 1.29 (1.04–1.59) | 0.022 |
| **Household wealth index (Ref: Poorest)** | | | | |
| Poorer | 0.88 (0.69–1.13) | 0.324 | 0.89 (0.69–1.14) | 0.340 |
| Middle | 0.75 (0.57–1.00) | 0.051 | 0.75 (0.57–1.00) | 0.052 |
| Richer | 0.69 (0.52–0.91) | 0.010 | 0.69 (0.52–0.92) | 0.012 |
| Richest | 0.55 (0.38–0.79) | 0.001 | 0.56 (0.38–0.81) | 0.002 |
| **Source of drinking water (Ref: Unimproved)** | | | | |
| Improved water source | 0.97 (0.80–1.17) | 0.733 | 0.96 (0.79–1.17) | 0.694 |
| **Sanitation facility (Ref: Unimproved)** | | | | |
| Improved sanitation | 0.94 (0.74–1.19) | 0.598 | 0.93 (0.74–1.18) | 0.564 |
| **Place of residence (Ref: Urban)** | | | | |
| Rural | 1.05 (0.88–1.25) | 0.586 | 1.05 (0.88–1.25) | 0.575 |
| **Département (Ref: Littoral)** | | | | |
| Alibori | 1.14 (0.71–1.81) | 0.591 | 1.15 (0.72–1.84) | 0.546 |
| Atacora | 1.10 (0.69–1.74) | 0.699 | 1.11 (0.70–1.77) | 0.650 |
| Atlantique | 1.06 (0.68–1.64) | 0.804 | 1.07 (0.69–1.66) | 0.747 |
| Borgou | 1.02 (0.66–1.56) | 0.930 | 0.93 (0.65–1.34) | 0.707 |
| Collines | 0.74 (0.46–1.20) | 0.226 | 0.76 (0.47–1.23) | 0.263 |
| Couffo | 1.24 (0.76–2.01) | 0.391 | 1.25 (0.77–2.02) | 0.367 |
| Donga | 0.79 (0.48–1.31) | 0.358 | 0.79 (0.48–1.30) | 0.346 |
| Mono | 1.04 (0.61–1.75) | 0.897 | 1.07 (0.63–1.80) | 0.810 |
| Ouémé | 1.18 (0.76–1.84) | 0.465 | 1.20 (0.77–1.86) | 0.424 |
| Plateau | 1.04 (0.67–1.63) | 0.857 | 1.05 (0.68–1.64) | 0.815 |
| Zou | 1.05 (0.68–1.62) | 0.824 | 1.07 (0.70–1.65) | 0.749 |
| **Model specification diagnostics** | | | | |
| *Goodness-of-fit test (p-value)* | 0.1795 | | 0.1973 | |
| *AIC (non-survey model)* | 5544.51 | | 5542.48 | |
| *BIC (non-survey model)* | 5802.55 | | 5832.77 | |
| *LRT: Model 2A vs Model 1A (p-value)* | — | | χ²(5) = 12.03; p = 0.034 | |
| *Mean VIF* | 2.00 | | 3.00 | |

Note: Survey-adjusted odds ratios (AOR) and 95% confidence intervals were estimated using svy. AIC and BIC values are derived from non-survey-weighted logistic regression models fitted to the same sample and are used for relative model comparison only. The likelihood-ratio test (LRT) compares Model 2A against Model 1A (nested). The sensitivity analysis sample differs from the main analytic sample (Table 3 of main manuscript, n = 10,153) by additionally restricting to children with complete data on maternal BMI and height, and by excluding children of currently pregnant mothers.

Abbreviations: AOR = Adjusted odds ratio; CI = Confidence interval; PBI = Preceding birth interval; LRT = Likelihood-ratio test; AIC = Akaike Information Criterion; BIC = Bayesian Information Criterion; VIF = Variance inflation factor; — = Not applicable.

**S3 Table. Survey-adjusted logistic regression models for the association between preceding birth interval and stunting among children aged 6–23 months: core analysis sample (Model M23, n = 3,831) and sensitivity analysis sample (Model M23A, n = 1,890), Benin DHS.**

| **Variables** | **Model M23 — Core sample (n = 3,831)** | | **Model M23A — Sensitivity sample (n = 1,890)** | |
| --- | --- | --- | --- | --- |
|  | **AOR (95% CI)** | **p-value** | **AOR (95% CI)** | **p-value** |
| **Preceding birth interval (months) (Ref: < 24 months)** | | | | |
| *< 24 months (Ref) (Ref)* | — |  | — |  |
| 24–32 months | 0.86 (0.67–1.12) | 0.268 | 0.72 (0.47–1.10) | 0.129 |
| 33–44 months | 0.82 (0.61–1.09) | 0.172 | 0.77 (0.50–1.19) | 0.241 |
| 45–56 months | 0.62 (0.44–0.88) | 0.007 | 0.56 (0.34–0.94) | 0.027 |
| > 56 months | 0.71 (0.49–1.02) | 0.064 | 0.77 (0.43–1.38) | 0.374 |
| No preceding birth (first-born) | 1.06 (0.75–1.50) | 0.740 | 1.14 (0.70–1.87) | 0.599 |
| **Child sex (Ref: Male)** | | | | |
| Female | 0.57 (0.49–0.67) | <0.001 | 0.69 (0.54–0.87) | 0.002 |
| **Birth type (Ref: Single)** | | | | |
| Multiple | 4.36 (2.93–6.50) | <0.001 | 5.87 (3.41–10.10) | <0.001 |
| **Ever breastfed (Ref: No)** | | | | |
| Yes | 0.63 (0.38–1.05) | 0.078 | 1.26 (0.57–2.80) | 0.565 |
| **Maternal level of education (Ref: None)** | | | | |
| Primary | 0.91 (0.73–1.14) | 0.415 | 0.83 (0.61–1.13) | 0.239 |
| Secondary or above | 0.63 (0.48–0.81) | 0.001 | 0.62 (0.42–0.92) | 0.017 |
| **Maternal BMI (Ref: Underweight) — M23A only** | | | | |
| Normal (18.5–24.9) | — | — | 0.79 (0.52–1.20) | 0.272 |
| Overweight (25.0–29.9) | — | — | 0.68 (0.40–1.15) | 0.153 |
| Obese (≥ 30.0) | — | — | 0.56 (0.29–1.07) | 0.080 |
| **Maternal height (Ref: < 155 cm) — M23A only** | | | | |
| 155–159 cm | — | — | 0.66 (0.50–0.87) | 0.003 |
| 160–164 cm | — | — | 0.29 (0.21–0.41) | <0.001 |
| ≥ 165 cm | — | — | 0.30 (0.20–0.45) | <0.001 |
| **Number of children under 5 in household (Ref: 1)** | | | | |
| 2 | 1.11 (0.87–1.42) | 0.411 | 1.23 (0.85–1.77) | 0.277 |
| ≥ 3 | 1.19 (0.92–1.54) | 0.175 | 1.15 (0.78–1.71) | 0.479 |
| **Household wealth index (Ref: Poorest)** | | | | |
| Poorer | 0.96 (0.74–1.23) | 0.732 | 1.06 (0.71–1.59) | 0.771 |
| Middle | 0.87 (0.68–1.12) | 0.277 | 0.88 (0.58–1.32) | 0.527 |
| Richer | 0.81 (0.61–1.08) | 0.147 | 0.87 (0.54–1.40) | 0.568 |
| Richest | 0.62 (0.41–0.92) | 0.017 | 0.73 (0.40–1.31) | 0.291 |
| **Source of drinking water (Ref: Unimproved)** | | | | |
| Improved water source | 0.98 (0.81–1.19) | 0.852 | 0.90 (0.68–1.20) | 0.468 |
| **Sanitation facility (Ref: Unimproved)** | | | | |
| Improved sanitation | 0.87 (0.66–1.13) | 0.292 | 1.02 (0.70–1.48) | 0.932 |
| **Place of residence (Ref: Urban)** | | | | |
| Rural | 1.00 (0.83–1.19) | 0.957 | 0.98 (0.74–1.29) | 0.872 |
| **Département (Ref: Littoral)** | | | | |
| Alibori | 1.43 (0.86–2.38) | 0.163 | 1.56 (0.81–3.00) | 0.185 |
| Atacora | 1.24 (0.73–2.10) | 0.423 | 1.41 (0.72–2.75) | 0.321 |
| Atlantique | 1.27 (0.78–2.09) | 0.339 | 1.23 (0.67–2.26) | 0.504 |
| Borgou | 1.05 (0.66–1.69) | 0.823 | 0.92 (0.50–1.66) | 0.772 |
| Collines | 0.86 (0.51–1.47) | 0.581 | 1.23 (0.63–2.41) | 0.544 |
| Couffo | 1.31 (0.80–2.16) | 0.279 | 1.01 (0.51–2.01) | 0.978 |
| Donga | 0.84 (0.51–1.41) | 0.517 | 0.87 (0.47–1.61) | 0.658 |
| Mono | 0.79 (0.45–1.38) | 0.402 | 0.89 (0.43–1.84) | 0.745 |
| Ouémé | 1.04 (0.61–1.75) | 0.895 | 0.97 (0.50–1.88) | 0.920 |
| Plateau | 1.02 (0.59–1.74) | 0.954 | 0.73 (0.35–1.51) | 0.398 |
| Zou | 1.25 (0.77–2.01) | 0.369 | 1.14 (0.63–2.07) | 0.662 |
| **Model specification diagnostics** | | | | |
| *Goodness-of-fit test (p-value)* | 0.7615 | | 0.9736 | |
| *Mean VIF* | 2.05 | | 2.05 | |

Note: Survey-adjusted odds ratios (AOR) and 95% confidence intervals were estimated using svy. Model M23 was fitted on the core analysis sample restricted to children aged 6–23 months and includes covariates that were associated with stunting at the 20% significance level in bivariate analyses; maternal BMI and height were not available in this dataset. Model M23A was fitted on the sensitivity analysis sample (restricted to children with complete maternal anthropometric data) and additionally adjusts for maternal BMI and height. Rows labelled "M23A only" were not included in Model M23.

Abbreviations: AOR = Adjusted odds ratio; CI = Confidence interval; PBI = Preceding birth interval; VIF = Variance inflation factor; — = Not applicable or not included in model.

**S4 Table. Survey-adjusted logistic regression models for the association between preceding birth interval and stunting among children aged 24–59 months: core analysis sample (Model M59, n = 6,322) and sensitivity analysis sample (Model M59A, n = 2,790), Benin DHS.**

| **Variables** | **Model M59 — Core sample (n = 6,322)** | | **Model M59A — Sensitivity sample (n = 2,790)** | |
| --- | --- | --- | --- | --- |
|  | **AOR (95% CI)** | **p-value** | **AOR (95% CI)** | **p-value** |
| **Preceding birth interval (months) (Ref: < 24 months)** | | | | |
| *< 24 months (Ref) (Ref)* | — |  | — |  |
| 24–32 months | 0.92 (0.75–1.13) | 0.431 | 0.84 (0.63–1.11) | 0.222 |
| 33–44 months | 1.01 (0.83–1.24) | 0.894 | 1.02 (0.77–1.34) | 0.909 |
| 45–56 months | 0.63 (0.49–0.82) | <0.001 | 0.55 (0.37–0.81) | 0.002 |
| > 56 months | 0.65 (0.49–0.85) | 0.002 | 0.58 (0.40–0.85) | 0.006 |
| No preceding birth (first-born) | 0.74 (0.59–0.94) | 0.013 | 0.65 (0.46–0.92) | 0.015 |
| **Child sex (Ref: Male)** | | | | |
| Female | 0.90 (0.80–1.00) | 0.060 | 0.86 (0.73–1.02) | 0.074 |
| **Birth type (Ref: Single)** | | | | |
| Multiple | 2.00 (1.42–2.82) | <0.001 | 1.88 (1.14–3.09) | 0.013 |
| **Maternal age at birth (years) (Ref: 20–24)** | | | | |
| 25–34 | 0.73 (0.60–0.88) | 0.001 | 0.66 (0.49–0.89) | 0.007 |
| ≥ 35 | 0.61 (0.49–0.76) | <0.001 | 0.61 (0.43–0.86) | 0.005 |
| **Maternal level of education (Ref: None)** | | | | |
| Primary | 0.97 (0.83–1.14) | 0.715 | 0.84 (0.65–1.10) | 0.206 |
| Secondary or above | 0.78 (0.64–0.96) | 0.017 | 0.80 (0.60–1.08) | 0.146 |
| **Maternal working status (Ref: Not working)** | | | | |
| Working | 0.94 (0.80–1.12) | 0.499 | 0.96 (0.71–1.29) | 0.788 |
| **Maternal BMI (Ref: Thin < 18.5 kg/m²) — M59A only** | | | | |
| Normal (18.5–24.9) | — | — | 0.76 (0.56–1.05) | 0.094 |
| Overweight (25.0–29.9) | — | — | 0.56 (0.39–0.81) | 0.002 |
| Obese (≥ 30.0) | — | — | 0.39 (0.24–0.64) | <0.001 |
| **Maternal height (Ref: < 155 cm) — M59A only** | | | | |
| 155–159 cm | — | — | 0.71 (0.55–0.90) | 0.005 |
| 160–164 cm | — | — | 0.38 (0.29–0.49) | <0.001 |
| ≥ 165 cm | — | — | 0.38 (0.27–0.52) | <0.001 |
| **Number of children under 5 in household (Ref: 1)** | | | | |
| 2 | 1.02 (0.87–1.21) | 0.770 | 1.21 (0.93–1.59) | 0.158 |
| ≥ 3 | 1.12 (0.95–1.32) | 0.162 | 1.33 (1.03–1.71) | 0.032 |
| **Household wealth index (Ref: Poorest)** | | | | |
| Poorer | 0.84 (0.70–1.01) | 0.060 | 0.79 (0.59–1.06) | 0.122 |
| Middle | 0.63 (0.52–0.77) | <0.001 | 0.69 (0.49–0.97) | 0.034 |
| Richer | 0.49 (0.40–0.60) | <0.001 | 0.62 (0.46–0.85) | 0.003 |
| Richest | 0.30 (0.22–0.40) | <0.001 | 0.48 (0.32–0.74) | 0.001 |
| **Source of drinking water (Ref: Unimproved)** | | | | |
| Improved water source | 0.96 (0.84–1.11) | 0.596 | 1.04 (0.82–1.32) | 0.736 |
| **Sanitation facility (Ref: Unimproved)** | | | | |
| Improved sanitation | 0.90 (0.76–1.08) | 0.270 | 0.91 (0.70–1.19) | 0.490 |
| **Place of residence (Ref: Urban)** | | | | |
| Rural | 1.05 (0.90–1.23) | 0.543 | 1.11 (0.90–1.36) | 0.345 |
| **Département (Ref: Littoral)** | | | | |
| Alibori | 0.80 (0.53–1.21) | 0.288 | 1.00 (0.55–1.80) | 0.993 |
| Atacora | 0.77 (0.52–1.16) | 0.220 | 1.01 (0.59–1.74) | 0.967 |
| Atlantique | 0.99 (0.66–1.49) | 0.980 | 1.01 (0.59–1.73) | 0.958 |
| Borgou | 0.92 (0.62–1.37) | 0.677 | 1.16 (0.67–2.01) | 0.587 |
| Collines | 0.54 (0.35–0.83) | 0.005 | 0.58 (0.32–1.07) | 0.082 |
| Couffo | 1.14 (0.75–1.75) | 0.538 | 1.42 (0.75–2.69) | 0.277 |
| Donga | 0.63 (0.40–0.99) | 0.046 | 0.73 (0.38–1.39) | 0.339 |
| Mono | 0.87 (0.55–1.35) | 0.527 | 1.21 (0.66–2.24) | 0.532 |
| Ouémé | 1.11 (0.74–1.65) | 0.621 | 1.38 (0.78–2.44) | 0.262 |
| Plateau | 1.19 (0.77–1.84) | 0.440 | 1.30 (0.74–2.27) | 0.356 |
| Zou | 1.21 (0.82–1.80) | 0.339 | 1.05 (0.60–1.84) | 0.870 |
| **Model specification diagnostics** | | | | |
| *Goodness-of-fit test (p-value)* | 0.1205 | | 0.8172 | |
| *Mean VIF* | 2.07 | | 2.07 | |

Note: Survey-adjusted odds ratios (AOR) and 95% confidence intervals were estimated using svy. Model M59 was fitted on the core analysis sample restricted to children aged 24–59 months. Model M59A was fitted on the sensitivity analysis sample and additionally adjusts for maternal BMI and height. Breastfeeding was not included in either model as it was not significantly associated with stunting in the bivariate analyses among this age group. Rows labelled "M59A only" were not included in Model M59.

Abbreviations: AOR = Adjusted odds ratio; CI = Confidence interval; PBI = Preceding birth interval; VIF = Variance inflation factor; — = Not applicable or not included in model.
